# Characterisation of diabetes-associated adipose tissue dysfunction across the spectrum of body mass index

**DOI:** 10.64898/2026.01.26.26344859

**Authors:** Oliver I Brown, Derek R Magee, Michael Drozd, Marcella Conning-Rowland, Marilena Giannoudi, Aya Shouma, Alexander-Francisco Bruns, Natalie J Haywood, Lee D Roberts, Joanna Kalucka, Samuel D Relton, Mark T Kearney, Kathryn J Griffin, Richard M Cubbon

## Abstract

Diabetes mellitus (DM) and obesity frequently coexist. Both are associated with adipose dysfunction, yet the contribution of DM remains uncertain. Using bulk transcriptomics of subcutaneous and visceral adipose tissue (SAT and VAT, respectively), we show that DM is associated with shared and distinct patterns of differential gene expression in these depots. Gene ontology analysis of hits across depots highlighted extracellular matrix, inflammatory pathways, metabolism, axon guidance and endoplasmic reticulum stress. Histology revealed larger SAT adipocytes in people with DM, but only in the overweight category. Body mass index (BMI)-stratified transcriptomic analyses of SAT identified DM-associated hits present only in the overweight group. These were validated in plasma protein form using UK Biobank, informing our development of an adipose risk score that predicted incident DM in overweight people beyond a clinical risk score. Hence, molecular signatures of diabetic SAT can define high-risk adiposity, which may aid the targeting of clinical interventions.

## Introduction

DM and obesity cause substantial mortality and morbidity across the world, with cardiovascular disease accounting for a significant proportion of this (1, 2). Whilst significant advances in anti-obesity therapy have the potential to retard this phenomenon in affluent populations, epidemiological projections suggest a growing global burden in decades to come (3, 4). Despite DM and obesity often co-existing, there remain large gaps in our understanding of how they adversely interact. We recently demonstrated that they act additively to promote a range of cardiovascular events, but with normal weight DM being associated with a similar or greater cardiovascular risk than obesity without DM (5). Notably this analysis showed that markers of adiposity were more strongly associated with adverse cardiovascular phenotypes than other commonly used clinical biomarkers, such as blood pressure and serum cholesterol. The available literature suggest that DM adversely affects both SAT and VAT, with genes regulating metabolism, inflammation, and extracellular matrix often being identified as differentially expressed (6-9). However, the studies describing human tissue are limited by modest sample sizes that do not allow characterisation of diabetic adipose dysfunction across the spectrum of BMI. We set out to address this using data from the Gene and Tissue Expression (GTEx) project (10), which includes RNA-sequencing and histology of >100 people with DM and >400 controls without DM across the spectrum of BMI. In particular, we aimed to define the molecular signature of diabetic adipose tissue dysfunction in people with normal weight, overweight and obesity, contrasting findings in SAT and VAT, using these data to inform candidate biomarkers that we tested in >50,000 UK Biobank participants (11).

## Results

The characteristics of GTEx participants included in our analyses of SAT and VAT are presented in **Supplemental Table 1**. In brief, this shows that for both adipose depots, participants with DM were older, had higher BMI, longer ischaemic times before tissue biopsy, were less likely to be white, and had a greater prevalence of prior myocardial infarction and hypertension. The proportion of male participants was unrelated to DM status in both the SAT and VAT cohorts. Importantly, these and other technical factors were included as covariates in our transcriptional analyses. Analyses within normal weight, overweight and obese strata revealed similar findings, with the exception of BMI being similar in participants with and without DM, as expected.

### DM is associated with common and distinct alterations in SAT and VAT transcriptomes

Comparing the SAT bulk transcriptomes of 168 donors with DM against 478 donors without DM in the GTEx cohort, we identified 1140 DEGs with FDR<0.05 (**Figure 1a, Supplemental Table 2**). A gene ontology analysis revealed diverse hits with FDR<0.05 (**Figure 1b-c, Supplemental Table 3**), including terms related to extracellular matrix, inflammatory pathways and axon guidance. Comparing the VAT bulk transcriptomes of 129 donors with DM against 407 donors without DM in the GTEx cohort, we identified 314 DEGs with FDR<0.05 (**Figure 1d, Supplemental Table 4**). A gene ontology analysis of these revealed diverse hits with FDR<0.05 (**Figure 1e-f, Supplemental Table 5**), including terms related to endoplasmic reticulum stress, metabolism, extracellular matrix and axon guidance. Comparing DEGs from SAT and VAT, only 69 were common to both (**Figure 1g, Supplemental Table 6**), yielding gene ontology terms related to axon guidance, extracellular matrix and endoplasmic reticulum (**Figure 1h, Supplemental Table 7**). These data suggest that diabetes is associated with both shared and distinct differences in SAT and VAT function.

**Figure 1:**
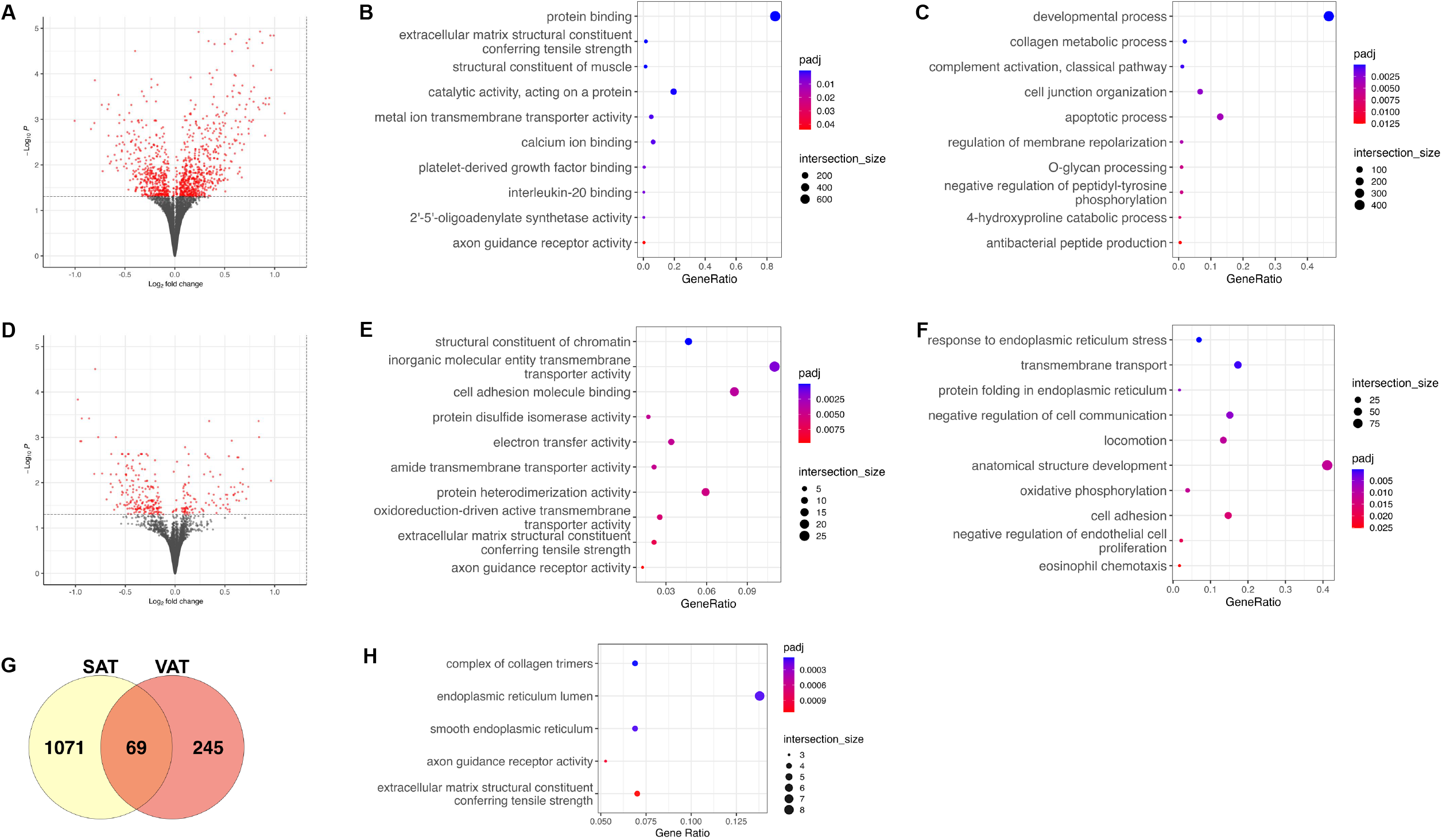
DM is associated with differential gene expression in SAT and VAT. **a**) Volcano plot illustrating differential expressed genes (DEGs) associated with diabetes mellitus (DM) in subcutaneous adipose tissue (SAT). **b**) Top 10 Molecular Function Gene Ontology (GO) terms attributed to SAT DEGs. **c**) Top 10 Biological Process GO terms attributed to SAT DEGs. **d**) Volcano plot illustrating DEGs associated with DM in visceral adipose tissue (VAT). **e**) Top 10 Molecular Function GO terms attributed to VAT DEGs. **f**) Top 10 Biological Process GO terms attributed to VAT DEGs. **g**) Venn diagram illustrating shared and distinct DEGs associated with DM in SAT and VAT. **h**) Top 5 GO terms attributed to DEGs shared by SAT and VAT.

### DM is associated with altered composition of SAT and VAT

Whilst single cell/nucleus sequencing technologies allow substantial characterisation of cellular heterogeneity in samples, deconvolution of bulk transcriptomic data can infer the high-level cellular composition of samples. Applying the CIBERSORTx algorithm, as we have previously published (12), we used the single nucleus data of Emont *et al* to define cell lineage expression signatures in human SAT and VAT (13), which we then applied to the GTEx cohort. In SAT, this revealed DM to be associated with lower abundance (p<0.05) of adipocytes and higher abundance of adipose stem and progenitor cells (ASPC), monocytes and macrophages (**Figure 2a**). In VAT, DM was associated with lower abundance of adipocytes and higher abundance of ASPCs and T lymphocytes (**Figure 2b**). These further support the notion of common and distinct impacts of DM on these depots.

**Figure 2:**
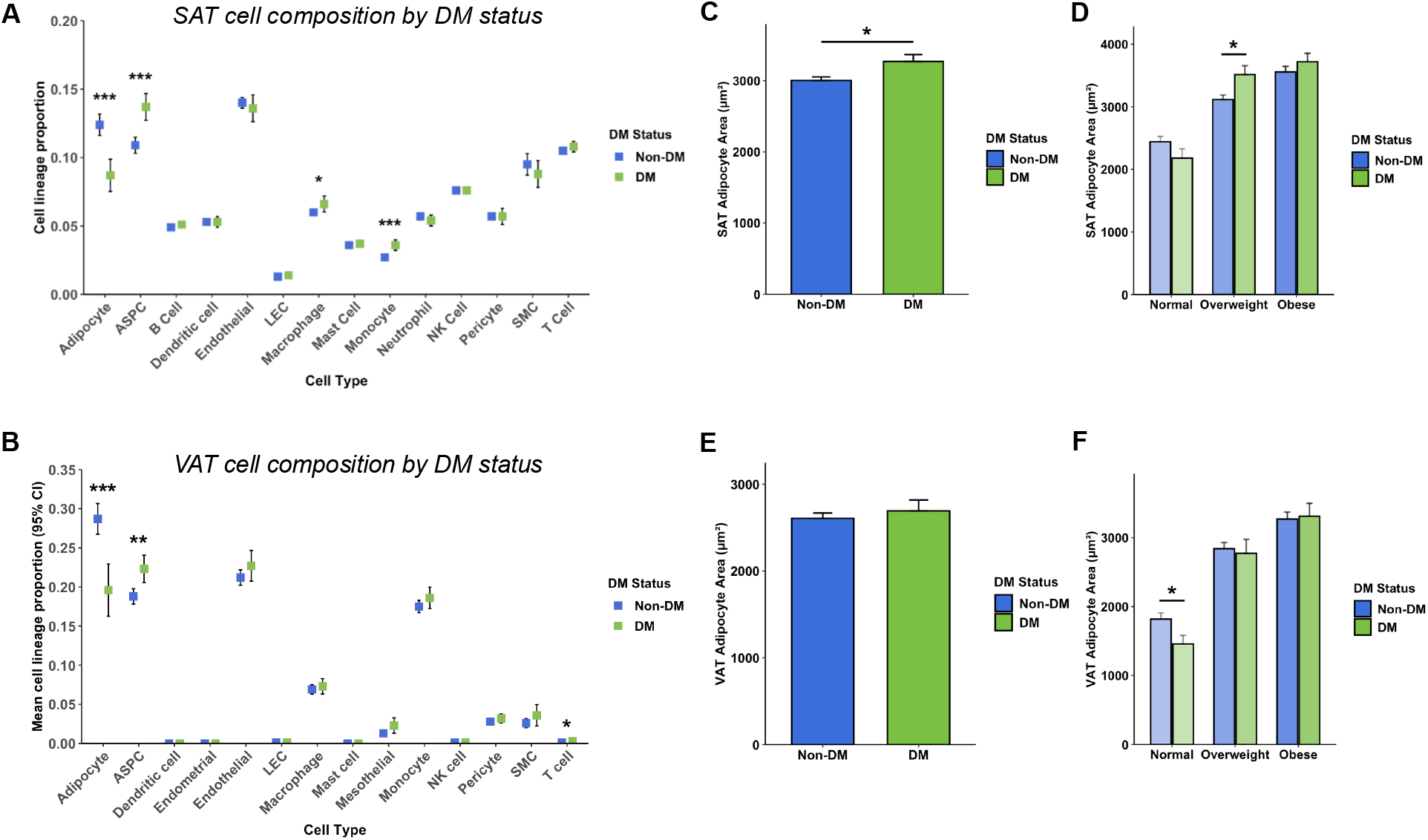
DM is associated with altered composition of SAT and VAT defined by RNA-seq deconvolution and histology. **a**) Cellular composition of subcutaneous adipose tissue (SAT) defined by RNA-seq deconvolution according to diabetes mellitus (DM) status. **b**) Cellular composition of visceral adipose tissue (VAT) defined by RNA-seq deconvolution according to DM status. **c**) Histological analysis of SAT adipocyte area according to DM status. **d**) Histological analysis of SAT adipocyte area according to DM status, stratified by body mass index (BMI) category. **e**) Histological analysis of VAT adipocyte area according to DM status. **f**) Histological analysis of VAT adipocyte area according to DM status, stratified by BMI category.

Next, we utilised the paired histology data in the GTEx resource to characterise the structural implications of diabetes, focussing on adipocyte size as a reproducible and validated marker of adipose structure, which is associated with adipose function. We defined median adipocyte area in 643 SAT samples (476 non-DM and 167 DM) and 533 VAT samples (405 non-DM and 128 DM). Analysis of SAT data revealed DM to be associated with larger adipocytes in the whole cohort (3003 vs. 3271μm^2^; p=0.014; **Figure 2c**). However, as SAT adipocyte area was moderately associated with BMI (R^2^=0.191; p<2×10^-16^), which is also associated with DM, we stratified our analysis according to clinical categories of BMI (normal, overweight and obese), revealing that only in the overweight group is DM associated with significantly larger adipocyte area (**Figure 2d**). Notably, higher SAT adipocyte area was positively correlated with CIBERSORTx inferred abundance of monocytes (R^2^=0.035; p=9.7×10^-7^) and neutrophils (R^2^=0.023; p=7.3×10^-5^). In contrast, analysis of VAT revealed no difference in adipocyte area between non-DM and DM groups (**Figure 2e**), although again there was important correlation between BMI and adipocyte area (R^2^=0.24; p<2×10^-16^). Therefore, we conducted BMI-stratified analyses, revealing DM to be associated with smaller adipocytes in the normal BMI group (1819 vs. 1458μm^2^; p=0.023; **Figure 2f**), although with no differences in the overweight and obese strata. Higher VAT adipocyte area was positively correlated with CIBERSORTx inferred abundance of endothelial cells (R^2^=0.027; p=9.4×10^-5^) and pericytes (R^2^=0.019; p=0.001). Hence, larger adipocytes are associated with distinct cellular changes in SAT versus VAT, with myeloid accumulation and vascular remodelling, respectively. Moreover, DM is associated with differing relationships between BMI and adipocyte area, which are depot specific, impacting the overweight range in SAT and the normal weight range in VAT. These data emphasise the complex association between DM and adipose tissue composition.

### Transcriptional characteristics of DM in SAT and VAT vary across the spectrum of BMI

Given the differential association of DM with adipocyte area across BMI groups, we redefined DEGs associated with DM in each of these BMI groups. For SAT, we identified 148 DEGs in people with normal weight (**Supplemental Table 8**; n=40 with DM vs. n=159 without DM), 425 DEGs in people with overweight (**Supplemental Table 9**; n= 64 vs. n=198), and 55 DEGs in people with obesity (**Supplemental Table 10**; n=64 vs. n=121). Notably, there was little overlap between DEGs in the 3 comparisons (**Figure 3a**), with only 11 shared DEGs in the normal and overweight groups (Cacna1b, Carmn, Dpys, Frk, Myh11, Nptxr, Pnck, Prima1, Osr1, Scube3 and Xkr4), 1 shared DEG in the normal and obese groups (Ptx3), and 3 shared DEGs in the overweight and obese groups (Caln, Ccdc13 and Chi3l1); all were directionally concordant. In the normal weight group, DM was associated with gene ontology terms relevant to the extracellular matrix and neuronal terms noted in the pan-BMI analysis (**Supplemental Table 11**). In the overweight group, gene ontology terms related to inflammation were the commonest also to be noted in the pan-BMI analysis (**Supplemental Table 12**). In the obese group, gene ontology terms related to inflammation and extracellular matrix were the commonest also to be noted in the pan-BMI analysis (**Supplemental Table 13**). BMI stratified analyses of our RNA-seq deconvolution data indicated DM was associated with lower adipocyte and higher monocyte abundance in the normal weight group; lower adipocyte and higher ASPC and monocyte abundance in the overweight group; lower adipocyte and higher ASPC, monocyte and B lymphocyte abundance in the obese group (**Figure 3b**).

**Figure 3:**
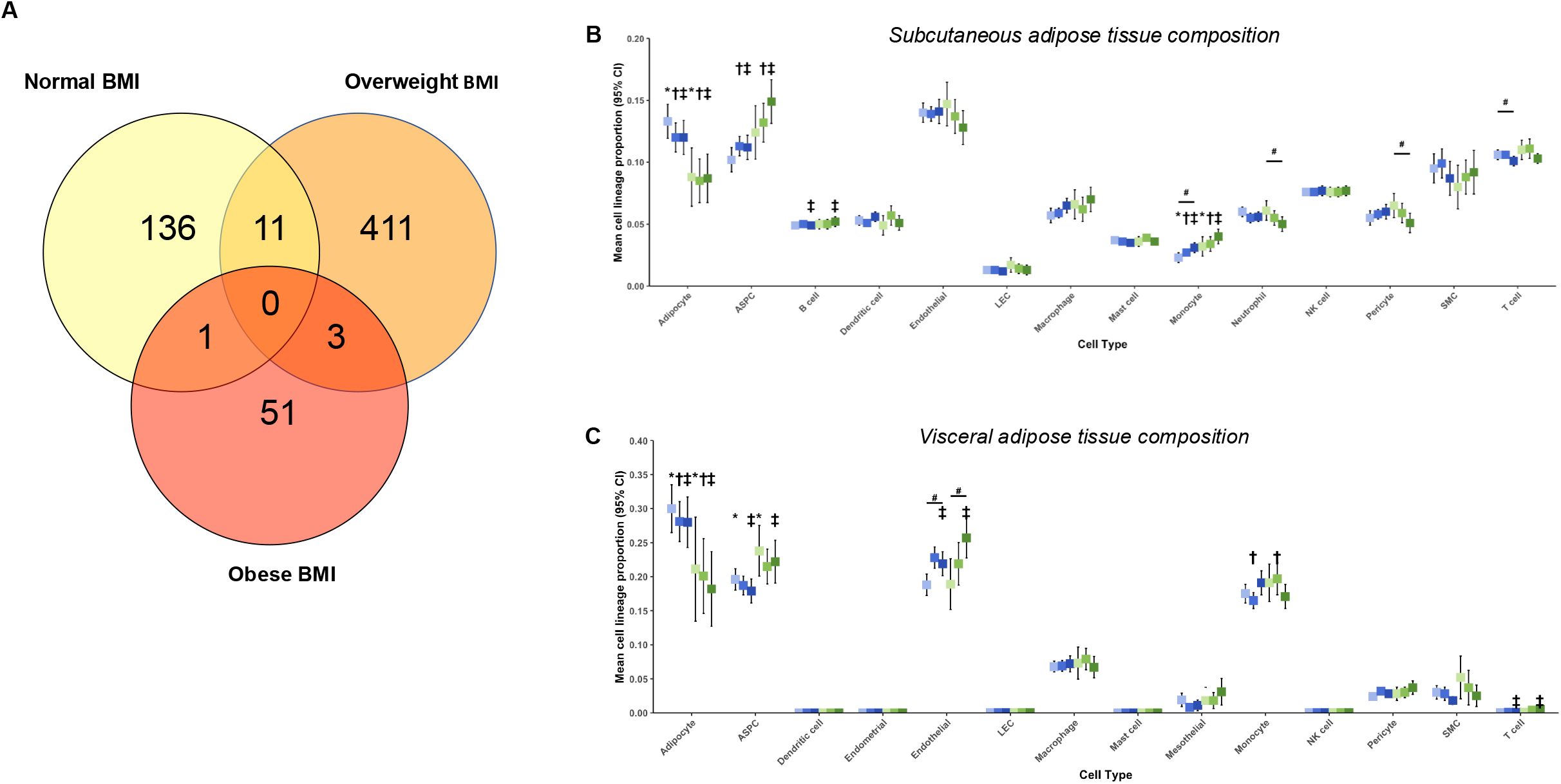
The adipose transcriptomic features of DM differ according to BMI category. **a**) Venn diagram illustrating shared and distinct differentially expressed genes (DEGs) associated with diabetes mellitus (DM) in subcutaneous adipose tissue (SAT) according to body mass index (BMI) category. **b**) Cellular composition of SAT defined by RNA-seq deconvolution according to DM status, stratified by BMI category. Blue denotes non-DM and green denotes DM; * denotes p<0.05 for non-DM vs. DM in the normal weight group; † denotes p<0.05 for non-DM vs. DM in the overweight group; ‡ denotes p<0.05 for non-DM vs. DM in the obese group; # denotes p<0.05 for normal weight vs. obese in the highlighted stratum with or without DM. **c**) Cellular composition of visceral adipose tissue defined by RNA-seq deconvolution according to DM status, stratified by BMI category. Annotation is identical to panel b.

For VAT, we identified 2 DEGs in people with normal weight (**Supplemental Table 14**; n=28 with DM vs. n=138 without DM), 23 DEGs in people with overweight (**Supplemental Table 15**; n= 51 vs. n=164), and 7 DEGs in people with obesity (**Supplemental Table 16**; n=64 vs. n=121), with no overlap between DEGs in the 3 comparisons. Given the small number of DEGs in the normal and obese groups, we only explored gene ontology analyses in the overweight group, revealing hits associated with inflammation and metabolism/transport (**Supplemental Table 17**), partially overlapping with the terms in the pan-BMI analysis. BMI stratified analyses of our RNA-seq deconvolution data indicated DM was associated with lower adipocyte and higher ASPC and monocyte abundance in the normal weight group; lower adipocyte and higher monocyte abundance in the overweight group; lower adipocyte and higher ASPC, endothelial and T lymphocyte abundance in the obese group (**Figure 3c**). Overall, these observations suggest that diabetes has both common and distinct implications for adipose function across the spectrum of BMI, especially in SAT. Moreover, even when stratified by BMI, diabetes remains associated with distinct abnormalities in SAT versus VAT.

### Validation of GTEx hits as plasma proteins in UK Biobank

Next, we sought to validate these adipose gene expression differences using an alternate methodology that facilitates development of clinical biomarkers. To do this, we used the UK Biobank resource to study DEG products in their plasma protein form (when present in the Olink 3072 panel), specifically testing for: 1) directionally concordant differential abundance of plasma proteins and differential expression of adipose gene expression in people with versus without DM; and 2) the association plasma proteins with diabetes and adipose tissue metrics. We included 52,743 participants of whom 3,263 (6.2%) had DM at recruitment. Among participants with DM, 387 (11.9%) were normal weight, 1,132 (34.7%) overweight, and 1,712 (53.4%) obese. In participants without DM, 15,822 (32.0%) were normal weight, 21,692 (43.8%) overweight, and 11,966 (24.2%) obese. For any given BMI class, participants with diabetes were more likely to be older, male, have had a prior myocardial infarction or hypertension, less likely to be white and have greater BMI (**Supplemental Table 18**). For SAT, of the 1140 DEGs, Olink plasma proteomic data were available for 212, of which 100 (47.2%) demonstrated directionally concordant differential expression at FDR<0.05 (**Supplemental Table 19**). Of these 100 plasma proteins, 63 were associated with BMI, 42 with abdominal SAT volume, 69 with VAT volume, 48 with hepatic fat %, 49 with muscle fat %, and 34 with HbA1c (a marker of glycaemia), all at FDR<0.05 (**Supplemental Table 16**). For VAT, of the 314 DEGs, Olink plasma proteomic data were available for 41, of which 13 (31.8%) demonstrated directionally concordant differential expression at FDR<0.05 (**Supplemental Table 19**). Of these 13 plasma proteins, 9 were associated with BMI, 6 with abdominal SAT volume, 9 with VAT volume, 7 with hepatic fat %, 6 with muscle fat %, and none with HbA1c, all at FDR<0.05 (**Supplemental Table 19**). Overall, these analyses provide evidence to support the external validity of a substantial proportion of hits found in our analyses of the GTEx cohort, along with supporting their potential role as circulating biomarkers.

### Derivation of an adipose risk score

Whilst obesity and DM (across the spectrum of BMI) are established targets for intensive intervention to improve metabolic health, overweight non-diabetic adults remain at risk of adverse sequelae due to adipose dysfunction. We reasoned that DEGs associated with DM in the overweight stratum of GTEx could inform the derivation of circulating biomarkers of early adipose dysfunction. To do this, we used the 425 DM-associated DEGs within the overweight group in SAT (**Figure 3a**), and applying the UK Biobank pipeline described above found 38 plasma proteins with directionally concordant differential expression to their gene counterparts (**Supplemental Table 20**). For proteins higher in people with DM, we dichotomised them into the upper quartile versus the lower 3 quartiles (reference group), and vice versa for proteins lower in people with DM, which we now refer to as in the high-risk quartile. For 36 of the 38 proteins (94.7%), non-diabetic participants in the high-risk quartile exhibited a higher incidence rate of DM (**Figure 4a**). When added to the Leicester Diabetes Risk Score (also known as the Diabetes UK risk score) (14), a tool used in the United Kingdom, the 38 dichotomised proteins improved model discrimination in predicting 10-year risk of DM in overweight people (AUC 0.736 [95% CI: 0.711–0.761] *vs* AUC 0.805 [95% CI: 0.783–0.827], p<0.001); **Figure 4b**); model discrimination also improved when all BMI categories were included (AUC 0.751 [95% CI: 0.731-0.772] *vs* AUC 0.805 [95% CI: 0.787-0.823], p<0,001; **Supplemental Figure 1**). Next, we applied LASSO regression to define the most informative proteins and established a set of 12 (ACP5, CDCP1, CHI3L1, EPHA1, ESM1, FBP1, GLA, IFI30, IL1RN, KYNU, MMP7 and PCSK9) that achieved a comparable increase in performance of the Leicester Diabetes Model (AUC 0.795 [95% CI: 0.773–0.817]; **Figure 4b**). People with a greater number of these proteins in the high-risk quartile exhibited higher rates of myocardial infarction, heart failure, atrial fibrillation and DM, in addition to greater hepatic fat percentage on magnetic resonance imaging (**Figures 4c-g**). These data suggest that a 12-protein blood panel, informed by the transcriptomic signature of diabetic adipose tissue, can identify people at risk of diabetes and wider sequelae of adipose dysfunction.

**Figure 4:**
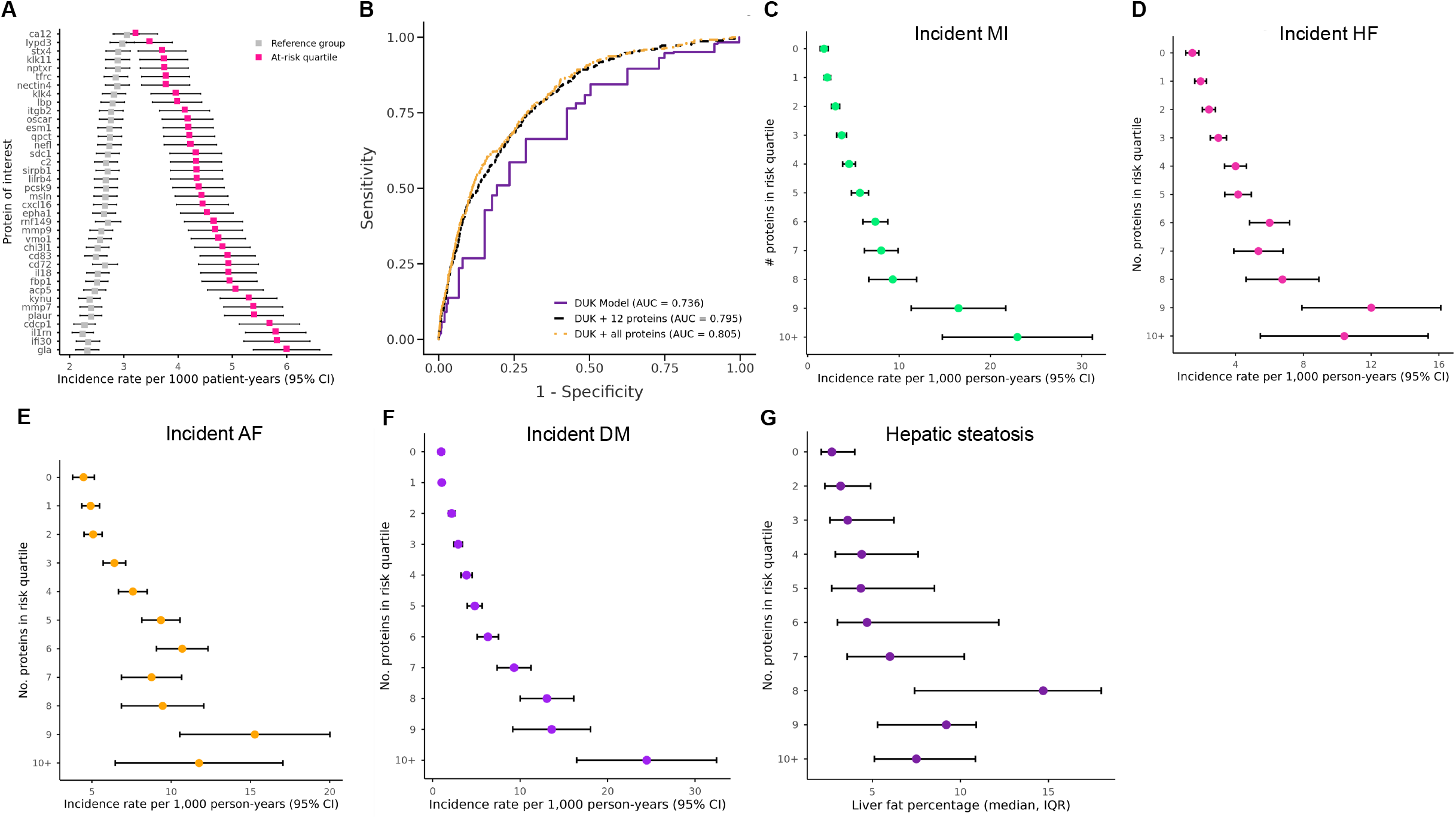
Plasma proteins defined from transcriptomic analysis of diabetic adipose predict incident DM and wider sequelae of adipose dysfunction in UK Biobank. **a**) Of 38 plasma proteins selected from differentially expressed genes associated with diabetes mellitus (DM) in overweight people, 36 are associated with increased incidence rate of DM when comparing people in the at-risk quartile vs the other 3 quartiles. **b**) The 38 proteins presented in panel a, and a subset of 12 defined with LASSO regression, improve the discriminative performance of the Leicester Diabetes Risk Model, illustrated using receiver operating characteristic (ROC) curves. **c-g**) Of the 12 proteins selected with LASSO regression, overweight participants with a greater number of proteins in the at-risk quartiles experience higher rates of myocardial infarction (**c**), heart failure (**d**), atrial fibrillation (**e**) and DM (**f**), along with greater hepatic fat percentage on magnetic resonance imaging (**g**).

## Discussion

The metabolic consequences of DM and obesity are often conflated, given their frequent coexistence. However, our analyses reveal a much more complex picture. We found that DM is associated with a distinct structural and functional adipose tissue phenotype, encompassing ECM changes, inflammation, altered metabolism and adipocyte hypertrophy, as has been previously reported. However, this phenotype was not uniform across the spectrum of BMI, and differed between SAT and VAT. Indeed, there was minimal overlap in DEGs associated with DM in normal, overweight or obese adipose tissue, whilst many enriched gene ontology terms were only present in part of the BMI spectrum. In SAT, the most pronounced diabetic phenotype was in the overweight range, leading us to particularly focus on this in our UK Biobank validation studies. These validated many adipose DEGs when assessed as circulating proteins, based on concordant differential abundance and association with adipose parameters. To address the clinical utility of this knowledge, we explored the value of a panel of circulating proteins in predicting incident DM in overweight people and found added value beyond a routine clinical risk score using metrics like age and waist-hip ratio. Moreover, these proteins were linked with features of adipose dysfunction, such as liver fat accumulation and cardiovascular events. This raises the potential of targeting intensive metabolic interventions, such as GLP1-RAs, to high-risk overweight individuals in clinical trials.

Whilst the design of our study cannot determine causal relationships, it is credible that DM could alter adipose phenotype (e.g. via its microvascular effects) and that adipose phenotypes could promote future DM (e.g. via lipodystrophy). Notably, DM was associated with distinct adipose phenotypes even in the normal weight range, with distinct gene expression profiles in both depots and smaller adipocytes in VAT. This aligns with other data showing that normal weight DM is associated with ectopic lipid deposition (implying a failure of adipose storage) and markedly increased cardiovascular events (5, 15). Our observation that DM is associated with the most marked SAT phenotype in the overweight range is particularly novel. Despite SAT adipocytes having comparable size in the normal and obese groups, their more marked hypertrophy in the DM overweight group may imply a failure of adipocyte hyperplasia. This possibility is supported by our CIBERSORTx deconvolution inferring a lower abundance of adipocytes and higher abundance of their stem/progenitor cells in diabetic SAT. Across the spectrum of BMI, DM was associated with changes in SAT metabolism and ECM, with normal weight also being linked to neuronal terms and overweight/obesity with inflammation terms. This concurs with many published transcriptomic analyses that have compared DM and non-DM donors, but without the BMI stratification we applied. Moreover, recent studies have shown that some of these biological phenomena (e.g. metabolic flexibility and ECM organisation) improve after weight loss interventions (16, 17), emphasising the potential value of biomarkers to define high risk adipose tissue.

An important part of our study was the identification of 12 proteins that independently predict incident DM beyond a clinically validated risk score. As a group, they provided a substantial increment in model discrimination beyond the clinical score, although independent replication of this using absolute protein concentrations is needed for this to proceed to use in clinical risk prediction. Notably, other studies have also used combinations of clinical and ‘omics biomarkers to achieve similar performance (18, 19). However, whilst their analyses applied an unbiased data-driven approach, our pipeline was informed by dysfunctional adipose tissue gene expression, optimising it to reflect adipose-mediated risk. This may allow improved targeting of intensive weight reduction strategies to individuals most likely to derive benefit, irrespective of BMI. Again, further validation studies are needed to assess this potential, potentially beginning with analysis of baseline proteomic data from clinical trials of weight loss interventions, such as GLP1-RAs. Regarding the biological insights provided by our 12 proteins, some have previously been linked to adipose tissue (dys)function. For example: ACP5 has been proposed as an adipokine in humans (20), its overexpression in mice leads to obesity (21), and *in vitro* studies suggest it regulates pre-adipocyte cell cycling (22); serum PCSK9 is higher in obese people and drops after bariatric surgery (23), knockout mice have greater adipocyte hypertrophy (24), and *in vitro* studies show it regulates adipocyte lipid uptake via regulation of CD36 degradation (25). Notably, we could find no published data linking adipose tissue (dys)function to other hits (including EHPA1, FBP1, and IFI30), so future study of these may elicit novel understanding of adipose biology. Finally, it is notable that single cell enrichment in the Tabula Sapiens SAT dataset of the Human Protein Atlas (https://www.proteinatlas.org) shows that 6 of the 12 proteins are enriched only in macrophages (ACP5, CDCP1, CHI3L1, FBP1, KYNU and MMP7). This suggests a future focus on SAT macrophages in people with diabetes could be particularly important.

It is also important to highlight some limitations of our study. First, the GTEx and UK Biobank cohorts largely focus on individuals between the ages of 40 to 70, most of whom are of white European origin. This means that caution should be applied when extrapolating our findings to other populations until further validation has been conducted. However, in many other ways the GTEx and UK Biobank cohorts are distinct and so the validation of many transcriptomic hits at circulating protein level adds confidence in their robustness. Second, our data cannot offer causal insight and so further work is needed to explore the biological meaning and therapeutic potential of hits. This does not detract from their potential as biomarkers, as we have demonstrated in our adipose risk score, although this will require external validation (for example, using the planned expansion of the UK Biobank plasma proteomics data). It will also be essential to define if high adipose risk defined by our score is modifiable by intensive metabolic intervention, for example by using GLP-1RAs beyond their current indications. Third, our data reflect a pooled population with type 1 and type 2 DM, most likely predominantly reflecting the latter due to its much greater prevalence; it will be important for future studies to conduct analyses restricted to these key subtypes.

In conclusion, DM is associated with a distinct phenotype in SAT and VAT, evident at both the structural and functional level, which is not uniform across the spectrum of BMI. Notably, we observed exaggerated hypertrophy of SAT adipocytes in overweight people with DM, along with features suggesting diminished adipocyte hyperplasia. A blood biomarker signature informed by the DM-associated DEGs in overweight SAT identifies overweight people with adverse metabolic health at high risk of future DM. This illustrates how knowledge of the adverse interactions between DM and adiposity can inform our understanding of metabolic health.

## Methods

### GTEx cohort

The Genotype-Tissue Expression (GTEx) project was established by the Broad Institute of MIT and Harvard in 2010 to create a repository of human genomic, transcriptomic and histology data to characterise multi-omic associations and disease mechanisms (10, 26). This includes RNA-sequencing from 54 tissues, collected post-mortem from organ donors with family consent, as described in detail on the GTEx project website (https://www.gtexportal.org). SAT was collected from the medial surface of the lower limb 2cm below the level of the patella and VAT from the omentum. Bulk RNA-sequencing raw count data were downloaded directly from the GTEx portal, using GTEx version 8 (phs000424.v8.p2 accessed July 2022). This includes metadata on donor age, gender, ischaemic time (between death and sample collection), and Hardy score (mode of death classification). Access to sensitive metadata related to comorbidities was obtained through an approved controlled-access application via dbGaP (https://www.ncbi.nlm.nih.gov/gap/) #32524 ‘Defining tissue-specific transcriptional profiles associated with diabetes’. RNA-sequencing and all required metadata were available for 646 SAT samples (478 without DM and 168 with DM) and 536 VAT samples (407 without DM and 129 with DM). Diagnoses of type 1 and type 2 DM were pooled since multiple donors were recorded as having both forms and detailed treatment data were not available to adjudicate. Body mass index (BMI) was used to categorise donors as normal weight (18.5–24.9 kg/m^2^), overweight (25–29.9 kg/m^2^), or obese (>30 kg/m^2^).

### Transcriptomics bioinformatic pipeline

Differential gene expression analysis was conducted as we have previously described (27), using R v.4.1.1 and the DESeq2 package v.1.36.0 (28). We sought DEGs associated with DM, with inclusion of the following covariates in the DESeq2 model: age in years (categorised as 20–29, 30–39, 40–49, 50–59, 60–69, and 70– 79); sex; race; ischaemic time in minutes (categorised as 0–299, 300–599, 600–899, 900–1199, and 1200–1499); Hardy score; BMI (categorised as normal, overweight or obese); medical history of hypertension; medical history of myocardial infarction; and RNA integrity score – RIN (categorised as 5.1–6, 6.1–7, 7.1–8, 8.1–9, and 9.1–10). Genes were filtered to include only those with greater than 10 read counts in at least the same number of samples as included in the DM subgroup. Effect size shrinkage using the apeglm method was applied for visualization and ranking of genes (29). False discovery rate (FDR)–adjusted P values produced by DESeq2 using the Benjamini–Hochberg method were used, with adjusted P<0.05 defined as statistically significant. Functional profiling of DEGs was performed using gProfiler (https://biit.cs.ut.ee/gprofiler/gost)(30), with significance threshold calculated using Benjamini–Hochberg FDR. Arising data were plotted using ggPlot2 v.4.0.1 (https://ggplot2.tidyverse.org).

Cell lineage proportions within VAT and SAT were estimated by deconvolution of GTEx bulk RNA-seq data using CIBERSORTx, as we have previously described in detail for other tissues (12). In brief, using human SAT and VAT single cell RNA-seq data published by Emont *et al* (13), 200 cells were selected at random to represent each cell lineage present in both depots, using data from donors with paired SAT and VAT single-cell RNA sequencing data, to generate single cell reference files for SAT and VAT. Each single cell reference file was uploaded to CIBERSORTx (https://cibersortx.stanford.edu/) to generate signature matrices and gene expression profiles to be used during deconvolution with default CIBERSORTx parameters and batch correction on. Cell lineage proportions were then imputed for each GTEx VAT and SAT bulk RNA-seq sample using their raw count data and the tissue-relevant signature matrix. Lineage proportions were compared between donors without versus with DM using t-tests, with some analyses stratified by BMI category as specified.

### Histology pipeline

We developed a multi-stage pipeline for adipocyte detection from digital whole slide images. The first stage is pixel level classification of pixels as either “white” or “non-white” (tissue) using a statistical model (histogram) trained on annotated examples of these two classes from on representative images from a local data set. In the second step, connected components analysis was used to identity connected white pixel areas (candidate adipocytes). The third step involved filtering the candidate regions by size (excluding areas <200um^2^ or >15000um^2^), elongation (excluding areas with elongation >2) based on bounding box, ratio of area/bounding box (exclude if ratio <0.5), and ratio of curve perimeter/circular area perimeter (excluded if ratio>2). The exclusions favour simple (cell shaped) shapes of the correct size over complex regions resulting from tears, or nearby cells that have been combined into a single region due to tissue damage. White background regions are also excluded these criteria. Visually, the method detected the majority of adipocytes, while including very few non-adipocytes (tears, background, multi-cell regions, background). The method was implemented in C++ using Microsoft Visual Studio (Microsoft, Redmond, USA), and the Open CV library (https://opencv.org), as an add-on method to the HeteroGenius-MIM digital pathology system (HeteroGenius, Leeds, UK: https://www.heterogenius.co.uk). We calculated the median area of the included adipocytes as a representative statistic for each donor, repeated for SAT and VAT separately.

### UK Biobank (UKB) Study Population

UKB is a prospective observational cohort study of 502,462 participants aged 37–73 years recruited from 22 assessment centres across the United Kingdom (UK) between 2006 and 2010. It is an open access resource developed using UK government and biomedical research charity funding, which links wide-ranging phenotypic and health care record data. The UKB resource is open to all bona fide researchers. Full details of its design and conduct are available online (https://www.ukbiobank.ac.uk). UKB received ethical approval from the National Health Service (NHS) Research Ethics Service (11/NW/0382); we conducted this analysis under the UKB application ID: 148871. All participants provided written informed consent, and the research was conducted in line with the Declaration of Helsinki. The study was reported according to the Strengthening the Reporting of Observational Studies in Epidemiology (STROBE) statement. Only study participants with available proteomic data were used in our analyses.

### UKB Plasma proteomics data

The UKB Pharma Proteomics Project (or UKB-PPP) is a collaboration between UKB and thirteen biotechnology and pharmaceutical companies that measured 2,923 unique proteins in a subset of 53,029 UKB participants. Plasma proteins were measured using the antibody based Olink Explore 3072 Proximity Extension Assay technology. Detailed methodology, quality control procedures and validation are described elsewhere (31). Protein concentrations were quantified as Normalised Protein eXpression (NPX) values (Olink’s arbitrary unit, expressed in a log_2_ scale) for each protein per participant. Values below the limit of detection were classified as missing. In the present analyses we used only protein measurements at baseline visit.

### Definitions of Diabetes, BMI, and Study Covariates in UKB analyses

Baseline sociodemographic characteristics, comorbidities, and medications were recorded by participants completing a touchscreen and nurse-led interview at study recruitment, as previously described (32). Data from the face-to-face nurse-led interview were used to ascertain baseline comorbidities and medications. Diabetes was classified as any diabetes (UKB field identifier 1220), type 1 diabetes mellitus (1222), type 2 diabetes mellitus (1223), diabetic eye disease (1276), diabetic neuropathy/ulcers (1468), and diabetic nephropathy (1607).

BMI was assessed using standing height and weight data collected at study recruitment. Participants classified as underweight were excluded. BMI category was adjusted for ethnicity in accordance with World Health Organization ethnicity-specific threshold recommendations: normal, BMI ≥18.5 kg/m^2^ to <25 kg/m^2^ or ≥18.5 kg/m^2^ to <23 kg/m^2^ if South Asian ethnicity; overweight, BMI ≥25 kg/m^2^ to <30 kg/m^2^ or ≥23 kg/m^2^ to <27.5 kg/m^2^ if South Asian ethnicity; and obese, BMI ≥30 kg/m^2^ or ≥27.5 kg/m^2^ if South Asian ethnicity (33). Definitions of other comorbidities at recruitment has previously been described in our published work (34).

### Assessment of Cardiometabolic Phenotype in UKB analyses

From 2014, all surviving participants were invited by e-mail and then postal mail to take part in multimodality imaging assessment, including abdominal MRI. Anthropometric measurements of body composition and measurements of serum lipids and biochemistry were collected at baseline. Responding participants were screened for eligibility for inclusion based on safety and tolerability criteria. All participants with metal implants in their body were excluded for safety and image quality concerns (35). Abdominal MRI was performed using a Siemens Aera 1.5T scanner (Syngo MR D13) (Siemens, Erlangen, Germany). The imaging protocol covered a 1.1m region from neck-to-knee region. A single 3D volume using an automated fat-water swap detection and correction procedure was calculated (36). Abdominal MRI data were available for 6,883 with proteomic data and included visceral adipose tissue volume (VAT) and abdominal subcutaneous adipose tissue volume (SAT). Liver fat percentage (data field 40061) was quantified using MRI-derived proton density fat fraction (PDFF), defined as the proportion of mobile proton signal attributable to fat relative to the total signal from fat and water in liver tissue. Weight and bioimpedance were measured using the Tanita BC418ma bioimpedance device (Tanita, Tokyo, Japan).

*Definition of Primary and Secondary Outcome Measures in UKB outcome analyses:* Our primary end point was incident DM, and our secondary end points were incident nonfatal myocardial infarction (MI), atrial fibrillation (AF) and heart failure (HF); all other analyses were exploratory. We used UKB-defined first disease occurrence fields to aid reproducibility of our definitions for incident DM (data field 130708), incident MI (data-field 131298), incident AF (data-field 131350) and HF (data-field 131354). Participants with a date of diagnosis before enrolment (therefore having pre-existing disease) were excluded. For incident disease outcomes, follow-up was limited to a maximum of ten years, with outcomes censored on 1st April 2024.

### Statistical analysis of UKB data

We analysed UKB data via the Research Access Platform secure cloud server (https://ukbiobank.dnanexus.com), using RStudio version 4.1.1. Analysis used the R suite ‘tidyverse’ (https://github.com/tidyverse), whilst figures were compiled using the ‘ggplot2’ sub-package. LASSO models and were constructed using ‘glmnet’ package (https://cran.r-project.org/web/packages/glmnet/index.html) and receiver operator curves (ROC) were generated using the ‘pROC’ package (https://cran.r-project.org/web/packages/pROC/index.html).

Continuous data are presented as median with 25th–75th centile. Categorical data are presented as count with percentage. Normality of distribution was checked using skewness and kurtosis tests; all continuous variables were found to be non-normally distributed. Differences between BMI categories within the diabetes or non-diabetes groups were assessed using Kruskal-Wallis H tests or χ2 test for continuous and categorical variables, respectively. Differences between the diabetes and non-diabetes groups within each BMI category were assessed using Mann-Whitney U tests. Correlation matrixes of Pearson model coefficients were used to assess for associations between proteins of interest and measures of metabolic phenotype. Proteins of interests from overweight participants without diabetes were split into quartiles derived from the UKB NPX value. The lowest or highest quartile (defined by over-representation in the DM group) was deemed the ‘at-risk’ quartile and the remaining three quartiles were grouped together at a reference group. Crude mortality rates were calculated per 1,000 person-years of follow-up for incident DM, nonfatal MI, AF and HF.

Penalised logistic regression using the least absolute shrinkage and selection operator (LASSO) was applied to identify circulating protein biomarkers associated with incident diabetes while limiting overfitting. Analyses were restricted to participants classified as overweight and conducted using complete-case data. LASSO models were fitted with ten-fold cross-validation to select the regularisation parameter (λ). Two values of λ were considered: the value minimising cross-validated prediction error (λ.min) and the largest value of λ within one standard error of the minimum (λ.1se). The λ.1se model was selected for primary inference as it yields a more parsimonious and stable set of predictors while maintaining predictive performance comparable to the minimum-error model. Proteins with non-zero coefficients at λ.1se were considered selected.

ROC curves were generated using binary logistic regression models for incident diabetes, comparing the discriminative performance of the Leicester Diabetes Risk Model to a protein panel and the Leicester Diabetes UK Risk score. The Leicester Diabetes UK Risk Model (https://riskscore.diabetes.org.uk) is a validated clinical risk score that estimates future type 2 DM risk based on basic demographic and anthropometric variables, including age, sex, ethnicity, body mass index, waist circumference, and family history of diabetes (14). DeLong’s test was used to define statistically significant differences in between model c-statistics.

All statistical tests were two-sided, and statistical significance was defined as P<0.05 after FDR adjustment. Missing data were not imputed.

## Data Availability

All data are available upon application to the GTEx portal (https://gtexportal.org/) and to UK Biobank (https://www.ukbiobank.ac.uk/)

https://www.ukbiobank.ac.uk/

## Acknowledgements

This research has been conducted using the UK Biobank Resource under Application Number 148871. This work uses data provided by patients and collected by the NHS as part of their care and support. This research used data assets made available by National Safe Haven as part of the Data and Connectivity National Core Study, led by Health Data Research UK in partnership with the Office for National Statistics and funded by UK Research and Innovation (research which commenced between 1st October 2020 – 31st March 2021 grant ref MC_PC_20029; 1st April 2021 -30th September 2022 grant ref MC_PC_20058).

## Data availability

The data underlying this article were accessed from the GTEx consortium (https://gtexportal.org) and UK Biobank (https://www.ukbiobank.ac.uk) and are available to other scientists after application to these organisations.

## Supplemental material

**Supplemental Table 1: Characteristics of GTEx cohort donors**. Characteristics of donors are presented separately for subcutaneous and visceral adipose tissue, both in whole cohort and after stratification by BMI categorised as normal, overweight or obese. Presented in Tab S1 of Supplemental_Table.xlsx

**Supplemental Table 2: SAT DEGs associated with DM**. Differentially expressed genes associated with diabetes mellitus in subcutaneous adipose tissue. Presented in Tab S2 of Supplemental_Table.xlsx

**Supplemental Table 3: GO terms relating to SAT DEGs associated with DM**. Gene ontology terms related to differentially expressed genes associated with diabetes mellitus in subcutaneous adipose tissue. Presented in Tab S3 of Supplemental_Table.xlsx

**Supplemental Table 4: VAT DEGs associated with DM**. Differentially expressed genes associated with diabetes mellitus in visceral adipose tissue. Presented in Tab S4 of Supplemental_Table.xlsx

**Supplemental Table 5: GO terms relating to VAT DEGs associated with DM**. Gene ontology terms related to differentially expressed genes associated with diabetes mellitus in visceral adipose tissue. Presented in Tab S5 of Supplemental_Table.xlsx

**Supplemental Table 6: Shared SAT and VAT DEGs associated with DM**. Differentially expressed genes associated with diabetes mellitus in both subcutaneous and visceral adipose tissue. Presented in Tab S6 of Supplemental_Table.xlsx

**Supplemental Table 7: GO terms relating to shared SAT and VAT DEGs associated with DM**. Gene ontology terms related to differentially expressed genes associated with diabetes mellitus in both subcutaneous and visceral adipose tissue. Presented in Tab S7 of Supplemental_Table.xlsx

**Supplemental Table 8: SAT DEGs associated with DM in normal weight subgroup**. Differentially expressed genes associated with diabetes mellitus in subcutaneous adipose tissue from normal weight subgroup. Presented in Tab S8 of Supplemental_Table.xlsx

**Supplemental Table 9: SAT DEGs associated with DM in overweight subgroup**. Differentially expressed genes associated with diabetes mellitus in subcutaneous adipose tissue from overweight subgroup. Presented in Tab S9 of Supplemental_Table.xlsx

**Supplemental Table 10: SAT DEGs associated with DM in obese subgroup**. Differentially expressed genes associated with diabetes mellitus in subcutaneous adipose tissue from obese subgroup. Presented in Tab S10 of Supplemental_Table.xlsx

**Supplemental Table 11: GO terms relating to SAT DEGs associated with DM in normal weight subgroup**. Gene ontology terms related to differentially expressed genes associated with diabetes mellitus in subcutaneous adipose tissue from normal weight subgroup. Presented in Tab S11 of Supplemental_Table.xlsx

**Supplemental Table 12: GO terms relating to SAT DEGs associated with DM in overweight subgroup**. Gene ontology terms related to differentially expressed genes associated with diabetes mellitus in subcutaneous adipose tissue from overweight subgroup. Presented in Tab S12 of Supplemental_Table.xlsx

**Supplemental Table 13: GO terms relating to SAT DEGs associated with DM in obese subgroup**. Gene ontology terms related to differentially expressed genes associated with diabetes mellitus in subcutaneous adipose tissue from obese subgroup. Presented in Tab S13 of Supplemental_Table.xlsx

**Supplemental Table 14: VAT DEGs associated with DM in normal weight subgroup**. Differentially expressed genes associated with diabetes mellitus in visceral adipose tissue from normal weight subgroup. Presented in Tab S14 of Supplemental_Table.xlsx

**Supplemental Table 15: VAT DEGs associated with DM in overweight subgroup**. Differentially expressed genes associated with diabetes mellitus in visceral adipose tissue from overweight subgroup. Presented in Tab S15 of Supplemental_Table.xlsx

**Supplemental Table 16: VAT DEGs associated with DM in obese subgroup**. Differentially expressed genes associated with diabetes mellitus in visceral adipose tissue from obese subgroup. Presented in Tab S16 of Supplemental_Table.xlsx

**Supplemental Table 17: GO terms relating to VAT DEGs associated with DM in overweight subgroup**. Gene ontology terms related to differentially expressed genes associated with diabetes mellitus in visceral adipose tissue from overweight subgroup. Presented in Tab S17 of Supplemental_Table.xlsx

**Supplemental Table 18: Characteristics of UK Biobank participants**. Characteristics of UK Biobank participants included in out analyses are presented in Tab S18 of Supplemental_Table.xlsx

**Supplemental Table 19: Associations between plasma proteins of interest, DM and adipose tissue phenotype in UK Biobank**. For selected plasma proteins of interest, proposed to inform about subcutaneous adipose tissue (SAT) or visceral adipose tissue (VAT) phenotype, data are presented on differential abundance in people with versus without diabetes. For proteins demonstrating concordant differential abundance to RNA-seq analyses in GTEx, linear associations amongst the whole cohort are presented for their z-transformed concentration against body mass index (BMI), abdominal SAT volume, VAT volume, hepatic fat percentage, muscle fat percentage, or glycated haemoglobin (HbA1c). Analyses are presented in Tab S19 of Supplemental_Table.xlsx

**Supplemental Table 20: Associations between plasma proteins of interest, DM and adipose tissue phenotype in UK Biobank within overweight cohort**. For selected plasma proteins of interest, proposed to inform about subcutaneous adipose tissue (SAT) or visceral adipose tissue (VAT) phenotype, data are presented on differential abundance in people with versus without diabetes for those within the overweight cohort. For proteins demonstrating concordant differential abundance to RNA-seq analyses in GTEx, linear associations amongst the whole cohort are presented for their z-transformed concentration against body mass index (BMI), abdominal SAT volume, VAT volume, hepatic fat percentage, muscle fat percentage, or glycated haemoglobin (HbA1c). Analyses are presented in Tab S19 of Supplemental_Table.xlsx

**Supplemental Figure 1:**
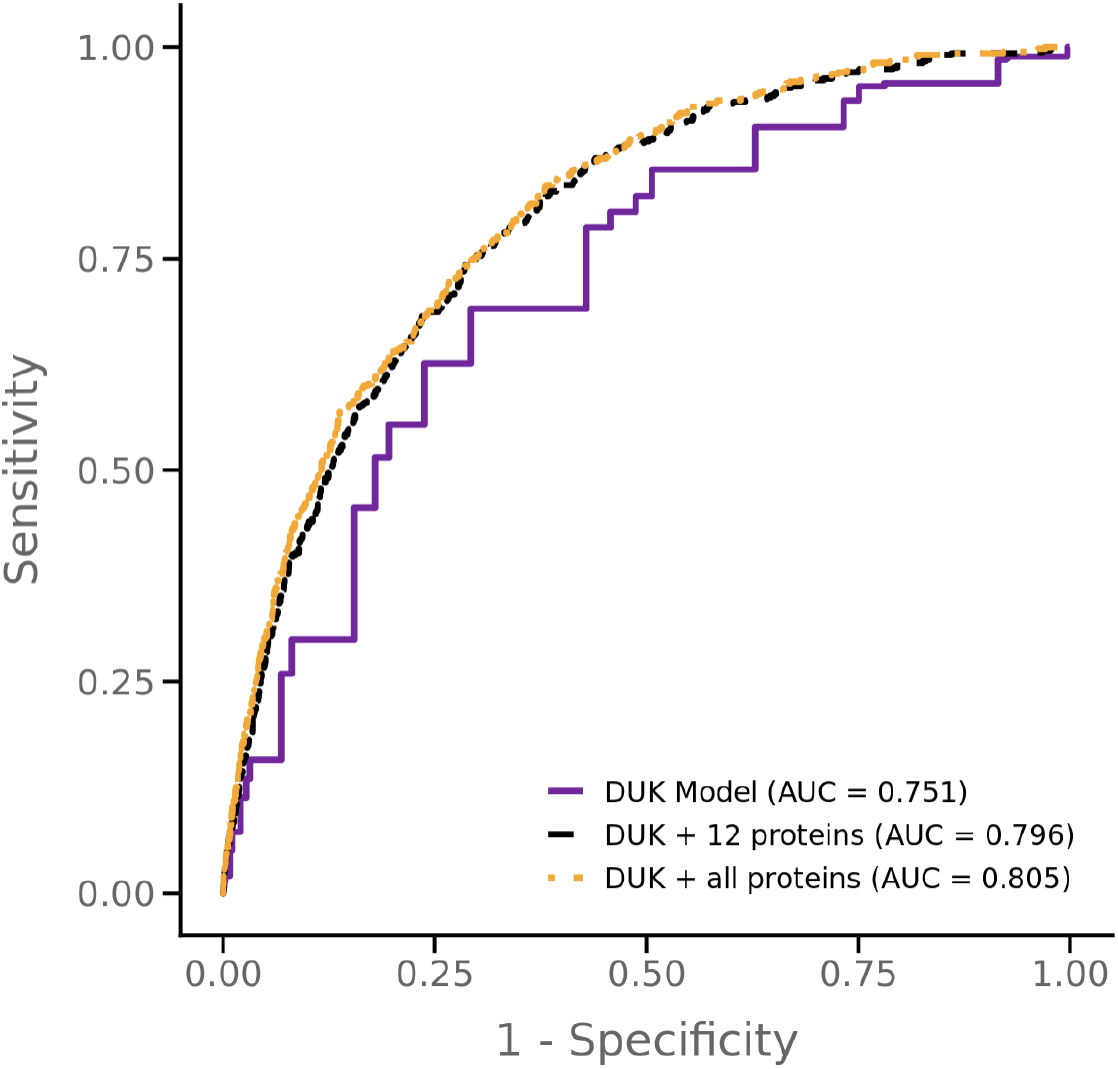
Performance of the Leicester Diabetes Model with or without plasma proteins of interest in UK Biobank. Receiver operating characteristic curves illustrating performance of the Leicester Diabetes Model alone, with the 38 proteins of interest, or the LASSO-selected 12 proteins of interest, in assessing risk of incident diabetes mellitus within 10 years.

